# Monitoring the proportion infected by SARS-CoV-2 from age-stratified hospitalisation and serological data

**DOI:** 10.1101/2021.01.11.21249435

**Authors:** Nathanaël Hozé, Juliette Paireau, Nathanaël Lapidus, Cécile Tran Kiem, Henrik Salje, Gianluca Severi, Mathilde Touvier, Marie Zins, Xavier de Lamballerie, Daniel Lévy-Bruhl, Fabrice Carrat, Simon Cauchemez

## Abstract

**Background:** Regional monitoring of the proportion infected by SARS-CoV-2 is important to guide local management of the epidemic, but is difficult in the absence of regular nationwide serosurveys.

**Methods:** We developed a method to reconstruct in real-time the proportion infected by SARS-CoV-2 and the proportion of infections being detected from the joint analysis of age-stratified seroprevalence, hospitalisation and case data. We applied our approach to the 13 French metropolitan regions.

**Findings:** We estimate that 5.7% [5.1%-6.4%] of adults in metropolitan France had been infected by SARS-CoV-2 by May 2020. This proportion remained stable until August and increased to 12.6% [11.2%-14.3%] by the end of November. With 23.8% [21.2%-26.8%] infected in the Paris region compared to 4.0% [3.5% - 4.6%] in Brittany, regional variations remained large (Coefficient of Variation CV: 0.53) although less so than in May (CV: 0.74). The proportion infected was twice higher (17.6% [13.4%-22.7%]) in 20-49 y.o. than in 50+ y.o (8.0% [5.7% - 11.5%]). Forty percent [33.7% - 45.4%] of infections in adults were detected in June-August compared to 55.7% [48.7% - 63.1%] in September-November. Our method correctly predicted seroprevalence in 11 regions in which only hospitalisation data were used.

**Interpretation:** In the absence of contemporary serosurvey, our real-time monitoring indicates that the proportion infected by SARS-CoV-2 may be above 20% in some French regions.

**Funding:** EU RECOVER, ANR, Fondation pour la Recherche Médicale, Inserm.

## Introduction

Less than a year after the emergence of SARS-CoV-2 and a first pandemic wave that has had devastating consequences, most European countries are now confronted with an intense second wave of SARS-CoV-2. In this context, the availability of up-to-date estimates of the proportion of the population that has been infected by SARS-CoV-2 and might potentially be immune in the different regions of a country is of critical importance to inform the local management of the epidemic. This information will become ever more important as the epidemic progresses and spatial heterogeneities in population immunity may keep on growing.

In many European countries, serological studies have provided estimates of the proportion of the population infected during the first pandemic wave. For example, it was estimated that about 4-5% of the population in metropolitan France had developed antibodies against SARS-CoV-2 by May 2020, with seroprevalences of the order of 10% in Grand Est and Ile-de-France, the two most affected regions (1–3). Since then, the virus has kept on circulating. Unfortunately, we lack contemporary estimates of seroprevalence that could capture the more recent regional evolution of the epidemic. This largely stems from the difficulty and cost to implement large-scale nationwide representative serosurveys at regular intervals. In this context, it is critical to develop methods that can track the proportion of the population that has been infected in the different regions from the joint analysis of existing seroprevalence data and other surveillance data that are more readily available in real time. Such monitoring is difficult to perform from the analysis of case data since testing practices changed substantially over time and space. Joint analysis of serological and death data across different countries has been used to reconstruct the proportion of infected individuals and allowed extrapolation to countries where serology was not available (4,5). However, such an approach may have difficulties to capture spread in younger age groups given low infection fatality ratios in these groups and may provide lagged estimates given the relatively long delays between infection and death.

Here, we present a method to reconstruct the proportion of the population infected by SARS-CoV-2 and the proportion of infections detected by surveillance from the joint analysis of age-stratified seroprevalence, hospitalisation and case data. The method is applied to metropolitan France and makes it possible to track in real time underlying SARS-CoV-2 infections by region and age group.

## Methods

### Estimation of age-specific infection hospitalisation ratio

Estimates of age-stratified infection hospitalisation ratios (IHR, i.e. the proportion of infected individuals in an age group that require hospitalisation) are derived from the joint analysis of hospitalisation and serological data documenting the impact of the first pandemic wave in Ile-de-France and Grand Est, the two regions of metropolitan France that were most affected. This calculation has been described elsewhere (6). In short, seroprevalence estimates were obtained from the SAPRIS study (1) gathering data from the large population-based French cohorts Constances, E3N-E4N, and NutriNet-Santé and the numbers of hospital admissions were obtained from the SI-VIC database, the national exhaustive inpatient surveillance system used during the pandemic. Since the median date of sample collection in the serosurvey was May 14, the IHR were obtained by dividing the cumulative numbers of hospital admissions up to May 6 (to account for estimated 11- and 19-day delays from infection to hospitalisation and seroconversion, respectively (7,8)) by the number of infected people estimated from the serosurvey. Patients from nursing homes, who were not part of the cohort target population, were excluded from the calculation.

### Sensitivity and specificity of the serological tests

Multiple imputation was used to infer the probability of infection among participants. In all participants, an EuroImmun IgG test against the S1 domain of the spike protein (Elisa-S1) was performed. When the Elisa-S1 optical density ratio was ≥0.7, two further tests (EuroImmun IgG test against Nucleocapsid protein and an in-house microneutralization assay to detect neutralizing anti-SARS-CoV2 antibodies) were performed. We assumed that participants with at least one positive test but no negative test were truly infected. Among the participants assumed truly infected, 82% (278/338) had three positive tests, 15% (52/338) had two positive tests and 2% (8/338) had one positive test.

Since the specificity was higher than 95% for each test independently (it was 100% for the neutralization assay (9)), the likelihood of two or three false positive tests in uninfected individuals could be considered negligible and the likelihood of one false positive test in uninfected individuals was very low and concerned very few participants. We therefore assumed the specificity to be 100%.

However, in this imputation model, an Elisa-S1 < 0.7 was sufficient to be classified as non-infected which may have been biased by the imperfect sensitivity of this serological method. We calculated the sensitivity of Elisa-S1 at this threshold (0.7) in participants with positive RT-PCR result in the cohort. We found that 91 participants had a positive SARS-CoV-2 RT-PCR less than 3 months before the serological test, among whom 76 had an Elisa-S1 ≥0.7, suggesting a sensitivity of the Elisa-S1 test at this threshold of 84% [75% - 90%]. This value was in line with the sensitivity reported at a threshold of 0.8 in an evaluation performed in SARS-CoV-2 PCR+ confirmed plasma donors (90.4% [84.4% - 94.7%] (10). We accounted for the imperfect sensitivity of the serological tests in the IHR. In our baseline scenario, we assumed the sensitivity of the test was 85% and considered 80%, 90% and 100% in sensitivity analyses.

### Reconstruction of the dynamics of infection

The curve of the daily number of infections was reconstructed from the daily number of hospital admissions and the distribution of the delay from infection to hospitalisation. For each age group, the number of infections was obtained as the deconvolution of the hospitalisation and the infection-to-hospitalisation delay distribution and divided by the IHR. The time-to-hospitalisation delay is discrete and parameterized with a gamma distribution with a mean of 11 days and standard deviation (sd) of 3.2 days. The deconvolution approach used a Richardson-Lucy scheme (11) that was adapted to account for right censoring in the hospitalisation curve (appendix p 1). The number of infections was reconstructed for all 13 regions of metropolitan France. Hospitalised individuals with missing age represented 0.7% of total hospitalisations (n=1480) and were not included in the study.

### External validation

The proportion of infected adults on May 11 was compared to the results of a national seroprevalence study (3) that was conducted in 12 regions of metropolitan France among those aged 15 y.o and over in May 2020.

### Estimation of the proportion of infections detected by surveillance

Dates of infections of confirmed cases were reconstructed with the same deconvolution approach using the national virological surveillance data (SI-DEP) and assuming the infection-to-detection delay has a gamma distribution of mean 8 days and sd 2.8 days, which accounts for an incubation period of 5.5 days (12) and a delay of 2.5 days to testing and reporting. Proportions of infections detected by surveillance were estimated over two periods (June-August and September-November), and were estimated as the ratio of the cumulative number of infections of cases over the cumulative number of infections obtained from the hospitalisations.

### Simulation study

We studied the accuracy of the deconvolution method at reconstructing the daily number of infections on simulated curves. Four infection curves were simulated to study various epidemiological scenarios (a gaussian epidemic, an increasing exponential, a decreasing exponential, a curve reconstructed from the hospitalisations in France between 1 September and 22 November). All but the first curve were right-censored.

The simulated curves of hospitalisations were then obtained by convolution with the infection-to-hospitalisation delay distribution and the daily infections were obtained using the Richardson-Lucy approach of (11) and our method. Comparison of the input and reconstructed infection curves are shown in appendix p 3.

## Results

Figure 1 presents the estimates of the seroprevalence (Figure 1A), the number of hospitalisations per 100,000 inhabitants (Figure 1B) and the IHR (Figure 1C) in the different age groups, in Ile-de-France and Grand Est. The seroprevalence was higher among the 40-49 y.o. (14% [12% - 17%]) and lower among the 70-89 y.o. (4% [3% - 7%]). The cumulative number of hospitalisations per 100,000 inhabitants and the IHR increased with age. The IHR increased from 0.46% [0.30% - 0.72%] in 20-29 y.o. to 21% [13% - 33%] in 70-89 y.o. Figure 1D shows the dynamics of hospitalisations by age group.

**Figure 1:**
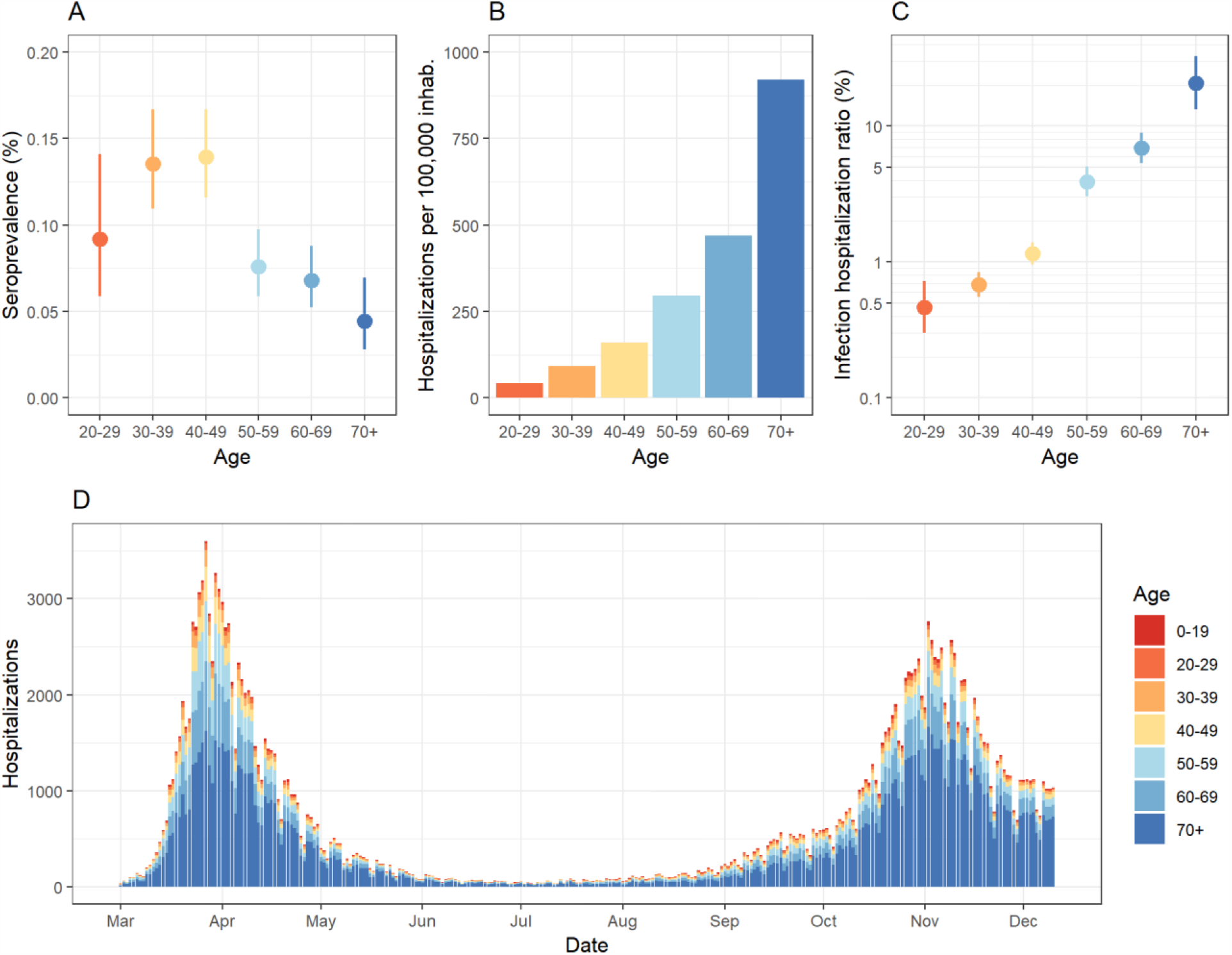
Description of data used for the study. (A) Estimates of seroprevalence by age group in Ile-de-France and Grand Est regions, in May-June 2020 (median date: 14 May). (B) Cumulative number of hospitalisations per 100,000 inhabitants, in Ile-de-France and Grand Est regions, from 1 March to 6 May 2020. (C) Estimates of infection hospitalisation ratio by age group (y-axis is in logarithmic scale). (D) Daily number of hospitalisations by age group in metropolitan France, from 1 March to 10 December 2020.

Our model is calibrated to serological data collected in two regions in May but can be used to reconstruct the seroprevalence and proportion infected in all regions and over time (Figure 2A-D). Consistent with national seroprevalence from our dataset for external validation (3), we estimate that 4.8% [4.3% - 5.4%] of adults were seropositive to SARS-CoV-2 in May 2020 in metropolitan France. Our regional estimates of seroprevalence in May are also consistent with those measured at the time, even in the 11 regions where only hospitalisation data informed our inference (Figure 2A). After correcting for the imperfect sensitivity of the serological assay, we find that 5.7% [5.1% - 6.4%] of the adult population had been infected by SARS-CoV-2 by May 2020 in metropolitan France (Figure 2B), with important regional variations (Figure 2C). The proportion infected remained stable during the summer 2020 and increased in September to reach 12.6% [11.2% - 14.3%] on November 30 (appendix p 4). On that date, the proportion infected was highest in Ile-de-France, Paris region (23.8% [21.2% - 26.8%]), followed by Provence-Alpes-Côte d’Azur (16.5% [14.4% - 18.9%]), Grand Est (15.1% [13.4% - 17.2%]), Auvergne-Rhône-Alpes (13.2% [11.6% - 15.0%]) and Hauts-de-France (12.4% [10.9% - 14.1%]) (Figure 2D). We find that the proportion infected is more homogeneous across regions in November than in May (coefficient of variation of 0.53 in November as opposed to 0.74 in May).

**Figure 2.**
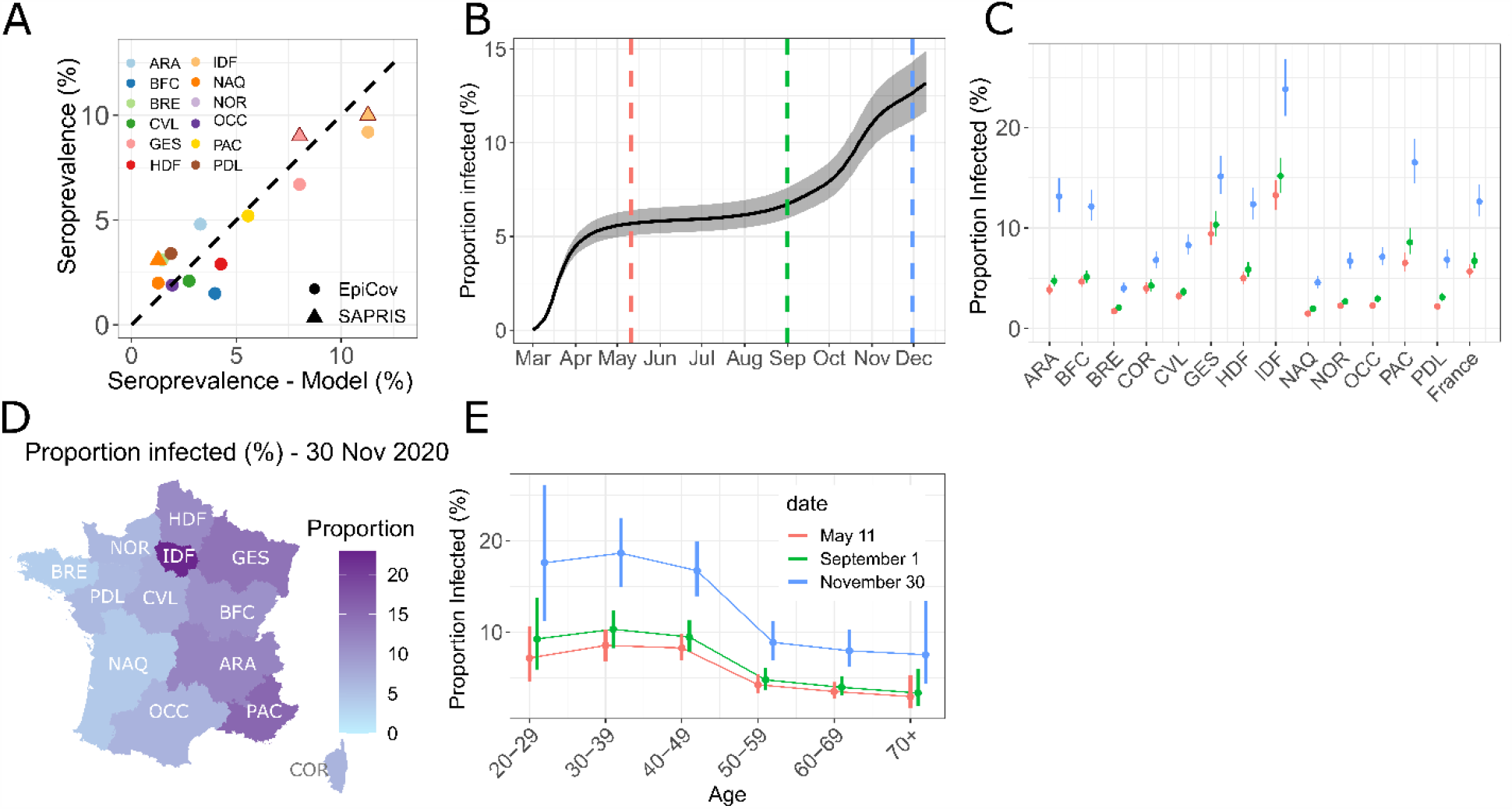
Reconstruction of the proportion infected in metropolitan France. (A) Scatter plot of the seroprevalence in regions estimated with our model on May 11 (x-axis) and in seroprevalence studies (y-axis) in May 2020. Data from SAPRIS serosurvey in IDF and GE (circled in red) were used to calibrate the model. (B) Proportion infected among adults in metropolitan France between 1 March and 10 December 2020. Timing of infection was reconstructed from the daily number of hospitalisation and the delay from infection to hospital admission. The grey area represents the 95% Confidence Interval. (C) Proportion infected in metropolitan France and in the 13 regions of metropolitan France on May 11 (red), September 1 (green) and November 30 (blue). (D) Geographical distribution of the proportion infected on November 30 2020. (E) Proportion infected by age group on May 11 (red), September 1 (green) and November 30 (blue). Abbreviations of regions name: ARA: Auvergne-Rhône-Alpes, BFC: Bourgogne-Franche-Comté, BRE: Bretagne, COR: Corse, CVL: Centre-Val de Loire, GES: Grand Est, HDF: Hauts-de-France, IDF: Île-de-France, NAQ: Nouvelle-Aquitaine, NOR: Normandie, OCC: Occitanie, PAC: Provence-Alpes-Côte d’Azur, PDL: Pays de la Loire.

The proportion infected was highest in those aged 20-49 years old (17.6% [13.4%-22.7%]) compared to 8.0 % [5.7% - 11.5%] in those aged 50+ y.o. (Figure 2E and appendix p 5). The same pattern by age was seen in most regions (Figure 3A), with the risk of infection in those aged 20-49 y.o. being 2-3 times higher than that in those aged 50+ y.o., depending on the region (Figure 3B).

**Figure 3.**
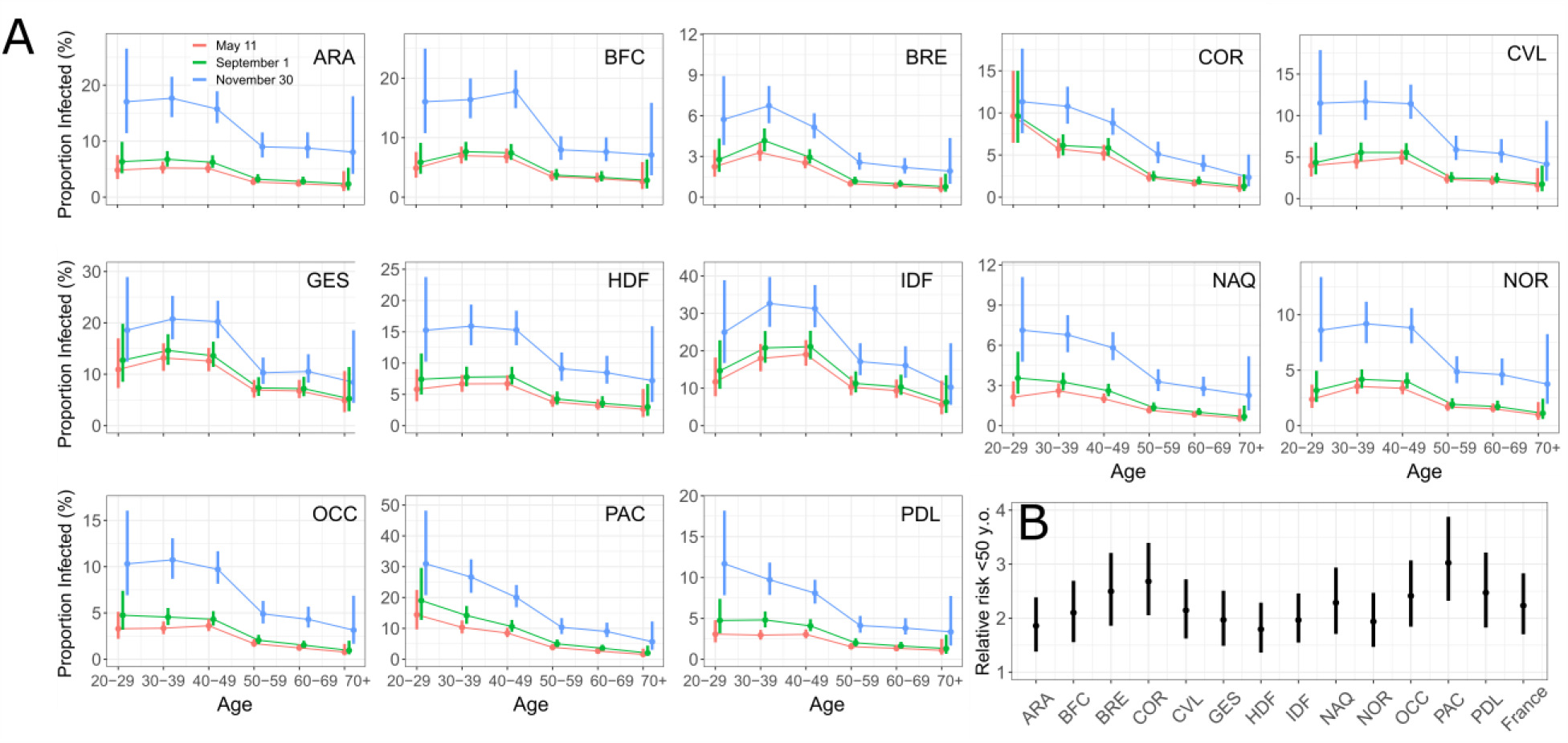
Proportion infected in the regions by age group and over time. (A) Estimates for the 13 regions of metropolitan France are shown, on May 11 (red), September 1 (green) and November 30 (blue). (B) Relative risk of infection of individuals under 50 y.o. compared to 50+ y.o.

We estimate that 53.6% [46.9% - 60.8%] of SARS-CoV-2 infections in the adult population were detected by surveillance between June and November 2020, with a probability of detection of 39.5% [33.7% - 45.4%] in June-August and 55.7% [48.7% - 63.1%] in September-November (p-value <0.001) (Figure 4 and appendix p 6). The probability of detection was higher in those aged >50 y.o. (68.6% [55.2% - 82.4%]) than in those aged 20-49 y.o (46.4% [38.8% - 54.2%]; p-value = 0.004) (Figure 4). These estimates are consistent with a simple analysis of the raw data: between June 1 and November 30, 120,000 adults were hospitalised and 1,800,000 cases were detected by surveillance, leading to a proportion detected of about 50% for an average estimated IHR of 3.3%.

**Figure 4.**
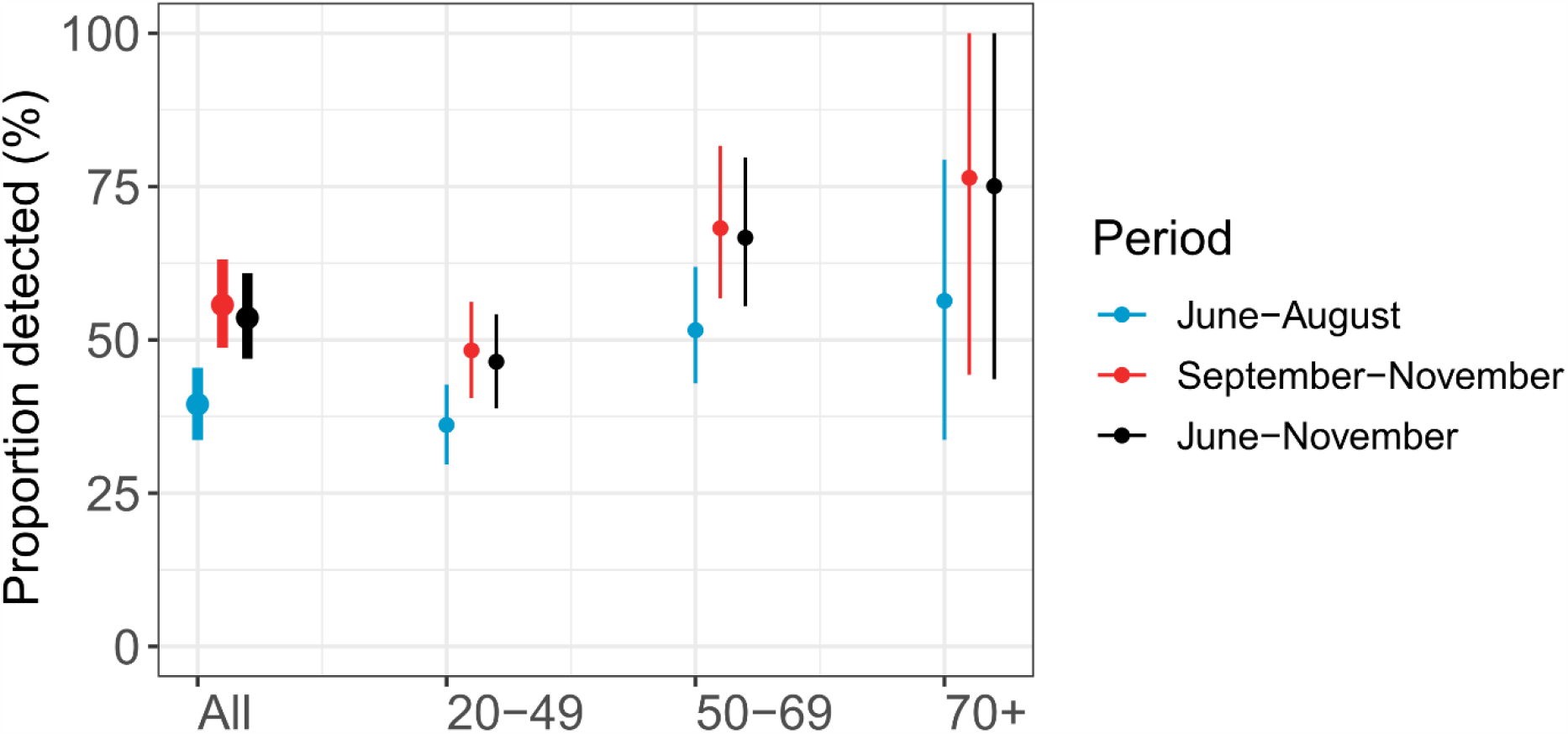
Proportion of infections detected by surveillance in June-August and in September-November.

In our baseline scenario, we assumed that the sensitivity of the serological test was 85%. In a sensitivity analysis, we find that estimates of the proportion infected by November 30 increased from 10.7% [9.5% - 12.2%] for a sensitivity of 100% to 13.4% [11.9% - 15.2%] for a sensitivity of 80% (Figure 5A) (p-value = 0.006). The proportion of infections that were detected varied from 50.4% [44.2% - 57.2%] for 80% sensitivity to 63.1% [55.2% - 71.6%] for 100% sensitivity (Figure 5B).

**Figure 5.**
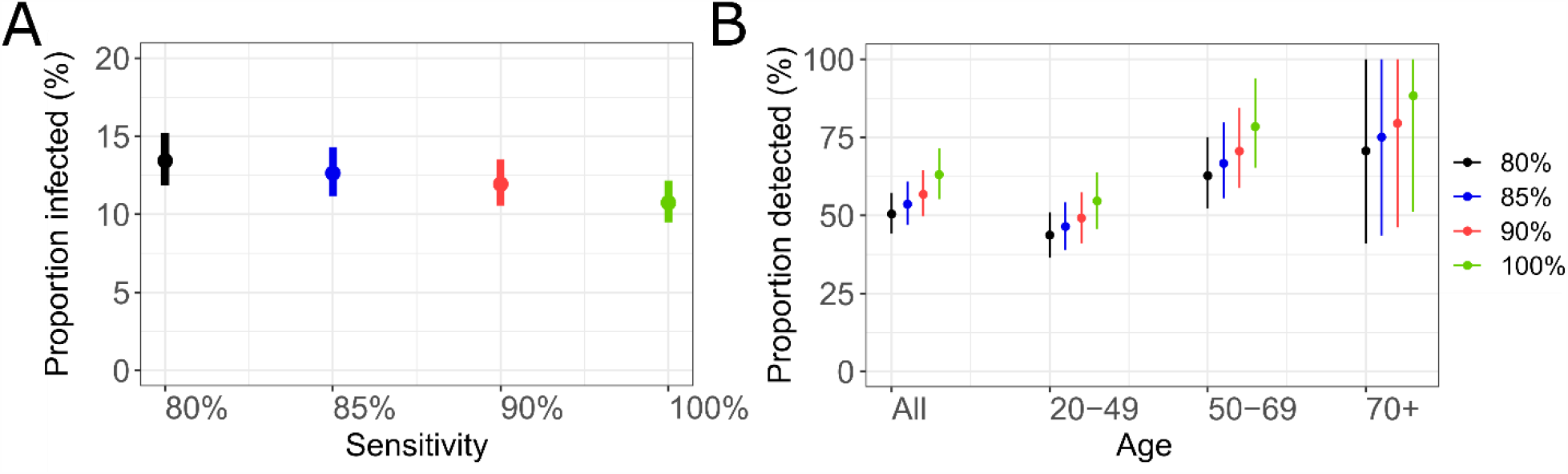
Sensitivity analysis. (A) Proportion infected on November 30, assuming 80%, 85%, 90%, 100% sensitivity of the serological tests. (B) Proportion of infections detected by surveillance between June and November, assuming 80%, 85%, 90%, 100% sensitivity of the serological tests. In our baseline analysis, we consider a sensitivity of the test of 85%.

## Discussion

We presented a method to reconstruct the proportion of the population infected by SARS-CoV-2 by region and age group from the joint analysis of readily available hospital surveillance data and existing serological surveys. The approach offers a simple way to track in real-time the number of infections in the population, which is challenging in the absence of regular, large-scale and representative serosurveys.

After accounting for the imperfect sensitivity of serology, we estimate that the proportion infected by SARS-CoV-2 in metropolitan France doubled from about 6% in May 2020 to about 13% at the end of November 2020, suggesting the two waves have relatively similar sizes. There are however important differences. First, the first wave occurred over a much shorter time period than the second wave that is still ongoing (Figure 2). Second, while the first wave was mostly concentrated in two regions, all regions were impacted by the second wave. As a consequence, the proportion infected was more homogeneous in November than in May 2020. However, substantial heterogeneities remain. For example, the proportion infected in Ile-de-France (Paris area) was about twice the national average. Overall, relatively similar patterns of infection by age were reconstructed in the different regions, with individuals aged 20-49 y.o. being at substantially higher risk (Figure 3B).

Assuming that those infected are immunized against reinfection, 24% immunity could contribute to slowing down the spread of the virus in Ile-de-France. Consider for example a situation in which control measures are such that, in a naive population, a case infects on average R0=1.6 persons. In such a scenario, we would expect the number of cases to double about every 10 days. With 20% immunity, the effective reproduction number would be reduced to Reff=0.8×1.6=1.3 leading to a substantially higher doubling time of about 18 days. However, given the very high transmissibility of SARS-CoV-2 in the absence of control measures (R0=3 (8)), 20% immunity would be insufficient to avoid a major sanitary crisis if all control measures were to be lifted. Indeed, in such a scenario, the effective reproduction number would be Reff=0.8×3=2.4, with the number of cases expected to double about every 5 days.

For the period between June and August 2020, we estimated that 39.5% [33.7% - 45.4%] of infections were detected, which is consistent with another modelling study (13) which reported a detection rate of 38% [35% - 44%] at the end of June. In our baseline scenario, we assumed a sensitivity of 85% for our assay, consistent with existing estimates (14). Higher sensitivities would lead to slightly lower estimates of the proportion infected and would inflate the proportion of infections being detected by surveillance at surprisingly high levels in some age groups (Figure 5).

Our estimates rely on the assumption that age-specific IHRs remained constant over time and across regions. However, it is possible that IHRs changed during the course of the pandemic, for example as a function of the stress on the healthcare system. Such variations could affect our estimates. For example, in a sensitivity analysis, we show that a 10-20% reduction in the IHR during the second wave would have limited impact on estimates of the proportion infected overall (13.3% [11.8% - 15.1%] for a 10% reduction and 14.1% [12.5% - 16.0%] for 20% reduction) (appendix p 3) but would reduce the proportion of infections being detected to 48.9% [42.8% - 55.4%] for 10% reduction in the IHR and to 44.0% [38.5% - 49.9%] for a 20% reduction) (appendix p 3). Our IHR estimates are calculated during the first pandemic wave and therefore constitute averages over a time period during which the stress on the healthcare system changed rapidly. IHRs may also vary with regional hospitalisation policies. For example, if there is higher propensity to hospitalize young adults in some regions, we might overestimate the proportion of infected in this age group and therefore in the overall population. However, while IHRs may vary by regions and with the stress on the healthcare system, we note that we were able to correctly predict the seroprevalence in the 13 regions of metropolitan France (Figure 2A) even though we only used IHRs estimates for Ile-de-France and Grand Est, the two regions that were the most affected during the first wave. Since our framework relies on the analysis of hospitalisation data and children have a very small probability of hospitalisation, our approach would likely generate large confidence intervals for that age group. We therefore decided to focus on adults.

Our approach makes it possible to estimate the number of persons that have been infected by SARS-CoV-2 since the start of the pandemic. It is believed that most persons infected by SARS-CoV-2 will acquire at least short term protection against reinfection (15). However, the duration of protection acquired through natural infection remains poorly characterized (16). In a scenario where there is waning of immunity and in the absence of vaccines, the estimated number of persons infected by SARS-CoV-2 will be an upper bound of the number of persons that are protected against infection. The interpretation of contemporary seroprevalence estimates may be equally challenging since it is unclear whether antibody decay following infection indicates a loss of protection. Therefore, in the long run, in the absence of vaccines, the proportion protected against SARS-CoV-2 may fall between the proportion seropositive estimated from seroprevalence studies and the proportion infected estimated with an approach such as the one presented here. Obviously, as vaccination campaigns are starting around the world, it will be essential to track the level of immunity acquired through vaccination.

We presented a simple framework to track the proportion infected in real-time from the joint analysis of age-stratified hospitalisation and serological data. Our approach should be easy to apply to other countries in which hospital surveillance and results of serosurveys are available.

## Data Availability

NA

## Contributors

NH, JP, NL, DLB, FC, SC designed and planned the study. NH, JP, NL, CTK, HS contributed to the statistical analysis. GS, MT, MZ, XdL, DLB, FC contributed to data collection. NH, JP, SC wrote the original draft. All authors critically edited the manuscript.

## Declaration of interests

Prof Fabrice Carrat reports personal fees from Imaxio and Sanofi, outside the submitted work. The other authors declare no competing interests.

## Acknowledgments

Support for this study was provided by ANR (Agence Nationale de la Recherche, #ANR-20-COVI-000, #ANR-10-COHO-06), Fondation pour la Recherche Médicale (#20RR052-00), Inserm (Institut National de la Santé et de la Recherche Médicale, #C20-26).

We acknowledge financial support from the Investissement d’Avenir program, the Laboratoire d’Excellence Integrative Biology of Emerging Infectious Diseases program (grant ANR-10-LABX-62-IBEID), Santé Publique France, the INCEPTION project (PIA/ANR-16-CONV-0005), and the European Union’s Horizon 2020 research and innovation program under grants 101003589 (RECOVER) and 874735 (VEO), AXA.

The CONSTANCES Cohort Study is supported by the Caisse Nationale d’Assurance Maladie (CNAM), the French Ministry of Health, the Ministry of Research, the Institut national de la santé et de la recherche médicale. CONSTANCES benefits from a grant from the French National Research Agency [grant number ANR-11-INBS-0002] and is also partly funded by MSD, AstraZeneca, Lundbeck and L’Oreal. The E3N-E4N cohort is supported by the following institutions: Ministère de l’Enseignement Supérieur, de la Recherche et de l’Innovation, INSERM, University Paris-Saclay, Gustave Roussy, the MGEN, and the French League Against Cancer. The NutriNet-Santé study is supported by the following public institutions: Ministère de la Santé, Santé Publique France, Institut National de la Santé et de la Recherche Médicale (INSERM), Institut National de la Recherche Agronomique (INRAE), Conservatoire National des Arts et Métiers (CNAM) and Sorbonne Paris Nord.The CEPH-Biobank is supported by the «Ministère de l’Enseignement Supérieur, de la Recherche et de l’Innovation».

