## Appendix for "Monitoring the proportion infected by SARS-CoV-2 from age-stratified hospitalisation and serological data"

**Reconstruction of infection curve with a deconvolution approach**

We used a deconvolution approach adapted from the method of Goldstein et al[1] to recover the unobserved curve of the daily number of infections from joint analysis of the daily number of hospitalisations and the infection-to-hospitalisation delay distribution.

Denote $(\lambda_1, \ldots, \lambda_N)$ and $(H_1, \ldots, H_N)$ the number of infections and hospitalisations, respectively, on days *1,...,N*. Denote $(d_1, \ldots, d_n)$ the infection-to-hospitalisation delay distribution, with $d_j$ the probability that an infected individual is admitted to hospital *j* days after infection, and $\sum_{j=1}^{n} d_i = 1$. We set *n = 40* and $d_i = 0$ if $i \leq 0$ or $i > n$. Denote *p* the probability that an infected individual is hospitalised.

The expected number of hospitalisations on day *i* is given by the convolution

$$H_i = p \sum_{j<i} d_{i-j}\lambda_j.$$

The deconvolution procedure reconstructs the daily number of infections iteratively by using an expectation maximization algorithm. The algorithm starts from an initial guess $({\lambda_1}^0, \ldots, {\lambda_N}^0)$ and the incidence vector is updated with the formula

$${\lambda_j}^{n+1} = \frac{{\lambda_j}^n}{q_j} \sum_{i>j} \frac{d_{i-j} H_i}{{H_i}^n},$$

where ${H_i}^n = \sum_{j<i} d_{i-j} {\lambda_j}^n$ is the expected number of hospitalisations that occur on day *i*, based on the vector of incidence at the *n*th iteration and $q_j = \sum_{-j+1 \leq i \leq \mathrm{N}-\mathrm{j}} d_i$ is a normalization factor that represents the probability that an hospitalisation resulting from an infection on day *j* will be observed during the interval *1...N*. The iteration is stopped when the normalized $\chi^2$ statistic, given by $\chi^2 = \frac{1}{N} \sum_i \frac{({H_i}^n - H_i)^2}{{H_i}^n}$, is below 1 for the first time.

In Goldstein et al[1] the deconvolution was applied to death curves of influenza epidemics and the initial vector of incidence was chosen to be the death curve shifted by nine days (mean time to death). This method was developed for an epidemic that had ended and is not directly applicable to the situation of a growing epidemic. Here, we adapted the method by choosing for the initial vector of incidence the (unshifted) hospitalisation curve multiplied by the probability of hospitalisation *p*.

We show in Figure S1 that this initialization vector allows reconstructing right-censored curves. We simulated a Gaussian epidemic (Fig S1A), an exponentially increasing infection curve (Fig S1B), exponentially decreasing (Fig S1C) and an infection curve obtained from the hospitalisation data in metropolitan France (Fig S1D). For each curve, we obtained the number of hospitalisations by convolution of the infection curve with the time-to-infection distribution and reconstructed the infection curve from the hospitalisation using the method of Goldstein et al. and with our method. For the hospitalisation data, we determined the number of infections using a deconvolution approach, obtained a second time the hospitalisations with a convolution of the infections, and then applied the method of Goldstein et al. and our method on the hospitalisations.

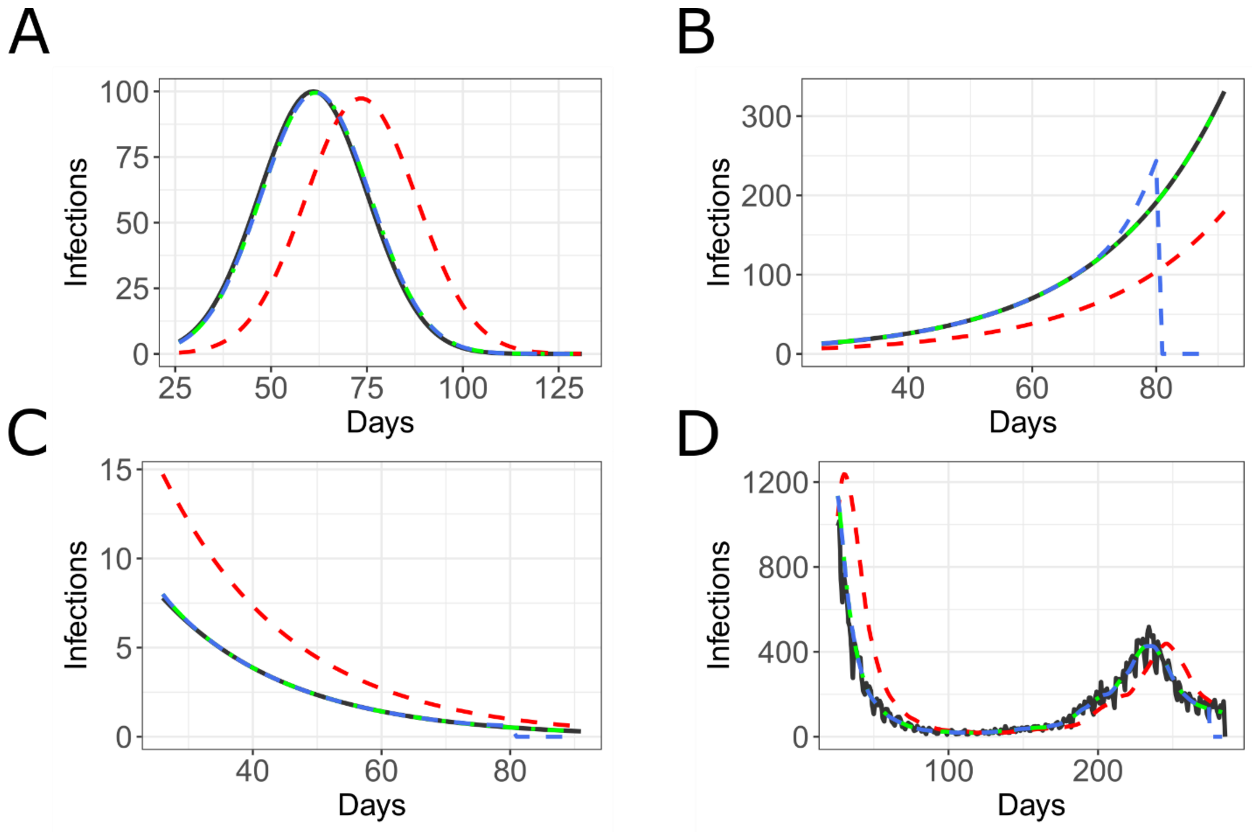


**Figure S1. Validation of the deconvolution reconstruction procedure.** The input (black) is plotted together with the hospitalisation curve (red) obtained by convolution with the time-to-hospitalisation delay distribution, and with the reconstruction from (Goldstein et al. 2009) (blue) and our adaptation (green). The method was tested on (A) a Gaussian epidemic, (B) an exponential increase, (C) an exponential decrease and (D) an infection curve obtained from the hospitalisation data in metropolitan France.

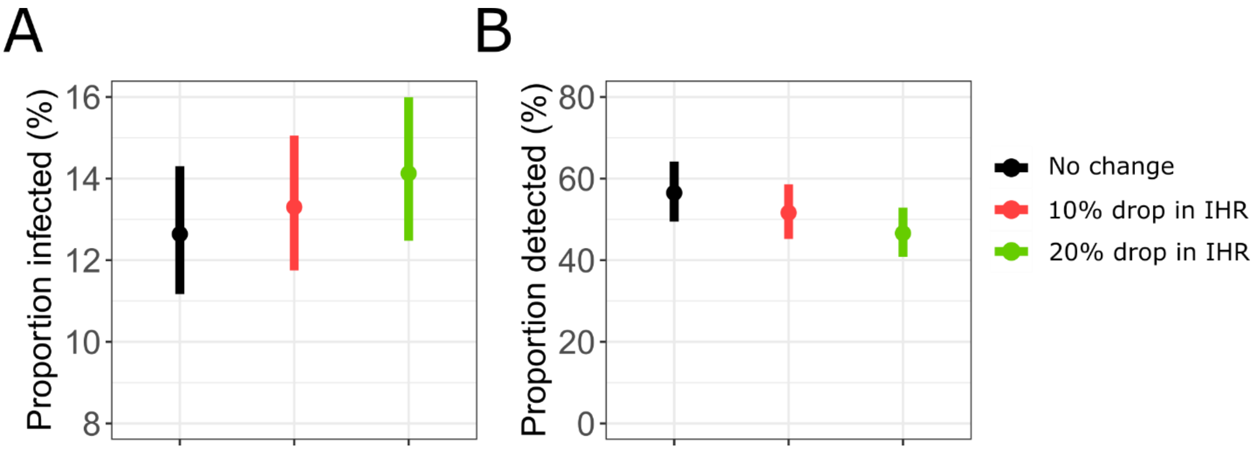


**Figure S2.** Impact of a reduction in IHR during the second wave on the proportion of infected (A) and the proportion of cases detected by surveillance (B).

**Table S1. Proportion infected among adults in the 13 regions of metropolitan France (mean and 95%CI).** Abbreviations: ARA: Auvergne-Rhône-Alpes, BFC: Bourgogne-Franche-Comté, BRE: Bretagne, COR: Corse, CVL: Centre-Val de Loire, GES: Grand Est, HDF: Hauts-de-France, IDF: Île-de-France, NAQ: Nouvelle-Aquitaine, NOR: Normandie, OCC: Occitanie, PAC: Provence-Alpes-Côte d'Azur, PDL: Pays de la Loire.

| | 11 May | 1 September | 30 November |
|---|---|---|---|
| ARA | 3.9% [3.4% - 4.4%] | 4.7% [4.2% - 5.4%] | 13.2% [11.5% - 15%] |
| BFC | 4.7% [4.1% - 5.3%] | 5.1% [4.6% - 5.8%] | 12.1% [10.7% - 13.8%] |
| BRE | 1.7% [1.5% - 1.9%] | 2.1% [1.9% - 2.4%] | 4% [3.5% - 4.6%] |
| COR | 4% [3.4% - 4.6%] | 4.3% [3.7% - 4.9%] | 6.8% [6% - 7.7%] |
| CVL | 3.2% [2.9% - 3.6%] | 3.7% [3.3% - 4.1%] | 8.3% [7.3% - 9.4%] |
| GES | 9.4% [8.3% - 10.6%] | 10.3% [9.1% - 11.7%] | 15.1% [13.4% - 17.2%] |
| HDF | 5% [4.4% - 5.7%] | 5.9% [5.2% - 6.6%] | 12.4% [10.9% - 14.1%] |
| IDF | 13.3% [11.8% - 14.8%] | 15.2% [13.5% - 17%] | 23.8% [21.2% - 26.8%] |
| NAQ | 1.5% [1.3% - 1.7%] | 2% [1.7% - 2.3%] | 4.6% [4% - 5.2%] |
| NOR | 2.3% [2% - 2.5%] | 2.7% [2.4% - 3%] | 6.7% [5.9% - 7.6%] |
| OCC | 2.3% [2% - 2.6%] | 3% [2.6% - 3.4%] | 7.1% [6.3% - 8.1%] |
| PAC | 6.5% [5.6% - 7.6%] | 8.6% [7.4% - 10%] | 16.5% [14.4% - 18.9%] |
| PDL | 2.2% [1.9% - 2.5%] | 3.1% [2.7% - 3.6%] | 6.9% [6% - 7.9%] |
| Metropolitan France | 5.7% [5.1% - 6.4%] | 6.7% [6% - 7.6%] | 12.6% [11.2% - 14.3%] |

**Table S2. Proportion infected among adults on November 30 (mean and 95%CI), by age group, in the different regions of metropolitan France.**

| | 20-29 | 30-39 | 40-49 | 50-59 | 60-69 | 70+ |
|---|---|---|---|---|---|---|
| ARA | 17% [26.5% - 11.4%] | 17.7% [21.5% - 14.3%] | 15.7% [18.9% - 13.2%] | 9% [11.6% - 7.1%] | 8.8% [11.6% - 7%] | 8.1% [18% - 4.2%] |
| BFC | 16% [25% - 10.8%] | 16.4% [20% - 13.3%] | 17.8% [21.4% - 14.9%] | 8% [10.2% - 6.3%] | 7.6% [10% - 6%] | 7.1% [15.9% - 3.7%] |
| BRE | 5.7% [8.9% - 3.8%] | 6.7% [8.2% - 5.4%] | 5.1% [6.2% - 4.3%] | 2.6% [3.3% - 2%] | 2.2% [2.9% - 1.7%] | 1.9% [4.4% - 1%] |
| COR | 11.3% [17.6% - 7.6%] | 10.8% [13.1% - 8.7%] | 8.8% [10.6% - 7.4%] | 5.1% [6.6% - 4%] | 3.8% [5% - 3%] | 2.4% [5.1% - 1.3%] |
| CVL | 11.5% [17.9% - 7.7%] | 11.7% [14.2% - 9.5%] | 11.4% [13.7% - 9.6%] | 5.9% [7.6% - 4.6%] | 5.4% [7.2% - 4.3%] | 4.2% [9.4% - 2.1%] |
| GES | 18.6% [28.9% - 12.4%] | 20.7% [25.2% - 16.8%] | 20.2% [24.3% - 17%] | 10.3% [13.2% - 8.1%] | 10.5% [13.9% - 8.4%] | 8.4% [18.6% - 4.4%] |
| HDF | 15.3% [23.8% - 10.2%] | 15.9% [19.3% - 12.9%] | 15.3% [18.4% - 12.8%] | 9.1% [11.7% - 7.2%] | 8.5% [11.2% - 6.7%] | 7.2% [15.9% - 3.8%] |
| IDF | 25% [38.9% - 16.7%] | 32.6% [39.7% - 26.4%] | 31.3% [37.6% - 26.3%] | 17.1% [22% - 13.5%] | 16.1% [21.2% - 12.8%] | 10.3% [22% - 5.5%] |
| NAQ | 7.1% [11.1% - 4.8%] | 6.8% [8.3% - 5.5%] | 5.8% [7% - 4.9%] | 3.3% [4.2% - 2.6%] | 2.8% [3.7% - 2.2%] | 2.3% [5.2% - 1.1%] |
| NOR | 8.6% [13.4% - 5.8%] | 9.2% [11.2% - 7.4%] | 8.8% [10.6% - 7.4%] | 4.9% [6.3% - 3.8%] | 4.6% [6.1% - 3.6%] | 3.7% [8.2% - 2%] |
| OCC | 10.3% [16.1% - 6.9%] | 10.7% [13.1% - 8.7%] | 9.7% [11.7% - 8.2%] | 4.9% [6.3% - 3.9%] | 4.3% [5.7% - 3.4%] | 3.1% [6.9% - 1.7%] |
| PAC | 30.9% [48.2% - 20.7%] | 26.6% [32.4% - 21.5%] | 20% [24.1% - 16.8%] | 10.4% [13.3% - 8.2%] | 9% [11.9% - 7.1%] | 5.7% [12.2% - 3.1%] |
| PDL | 11.7% [18.2% - 7.8%] | 9.7% [11.8% - 7.9%] | 8.1% [9.7% - 6.8%] | 4.1% [5.3% - 3.2%] | 3.8% [5% - 3%] | 3.3% [7.7% - 1.7%] |
| Metropolitan France | 17.6% [11.2% - 26.1%] | 18.6% [14.9% - 22.4%] | 16.7% [13.9% - 19.9%] | 8.9% [6.9% - 11.2%] | 8% [6.2% - 10.3%] | 7.5% [4.4% - 13.4%] |

**Table S3. Proportion of cases detected by surveillance, by age groups, from June to August, September to November, June to November.**

| Age | Period | Proportion detected |
|---|---|---|
| All | June-August | 39.5% [33.7% - 45.5%] |
| 20-49 | June-August | 36.1% [29.7% - 42.7%] |
| 50-69 | June-August | 51.6% [42.9% - 61.9%] |
| 70+ | June-August | 56.4% [33.7% - 79.4%] |
| All | September-November | 55.7% [48.7% - 63.1%] |
| 20-49 | September-November | 48.3% [40.5% - 56.2%] |
| 50-69 | September-November | 68.2% [56.7% - 81.6%] |
| 70+ | September-November | 76.4% [44.3% - 100%] |
| All | June-November | 53.6% [46.9% - 60.8%] |
| 20-49 | June-November | 46.4% [38.8% - 54.2%] |
| 50-69 | June-November | 66.7% [55.5% - 79.8%] |
| 70+ | June-November | 75.1% [43.6% - 100%] |
